# Risk stratification of hospitalized COVID-19 patients through comparative studies of laboratory results with influenza

**DOI:** 10.1101/2020.05.18.20101709

**Authors:** Yang Mei, Samuel E. Weinberg, Lihui Zhao, Adam Frink, Chao Qi, Amir Behdad, Peng Ji

**Author notes:** Equal contribution (Yang Mei, Samuel E. Weinberg). Correspondence to: Peng Ji, M.D., Ph.D., Department of Pathology, Feinberg School of Medicine, Northwestern University, 303 East Chicago Avenue, Chicago, IL 60611.

## Abstract

**Background:** The outbreak of coronavirus disease 2019 (COVID-19) in December 2019 overlaps with the flu season.

**Methods:** We compared clinical and laboratory results from 719 influenza and 973 COVID-19 patients from January to April 2020. We compiled laboratory results from the first 14 days of the hospitalized patients using parameters that are most significantly different between COVID-19 and influenza and hierarchically clustered COVID-19 patients based on these data. The clinical outcomes were compared among different clusters.

**Findings:** Temporal analyses of laboratory results revealed that compared to influenza, patients with COVID-19 exhibited a continued increase in the white blood cell count, rapid decline of hemoglobin, more rapid increase in blood urea nitrogen (BUN) and D-dimer, and higher level of alanine transaminase, C-reactive protein, ferritin, and fibrinogen. Using these results, we sub-classified the COVID-19 patients into 5 clusters through a hierarchical clustering analysis. We then reviewed the medical record of these patients and risk stratified them based on the clinical outcomes. The cluster with the highest risk showed 27·8% fatality, 94% ICU admission, 94% intubation, and 28% discharge rates compared to 0%, 38%, 22%, and 88% in the lowest risk cluster, respectively. Patients in the highest risk cluster had leukocytosis including neutrophilia and monocytosis, severe anemia, increased red blood cell distribution width, higher BUN, creatinine, D-dimer, alkaline phosphatase, bilirubin, and troponin.

**Interpretation:** There are significant differences in the clinical and laboratory courses between COVID-19 and influenza. Risk stratification in hospitalized COVID-19 patients using laboratory data could be useful to predict clinical outcomes and pathophysiology of these patients.

## Introduction

Coronavirus disease 2019 (COVID-19) rapidly disseminated in most countries. The causative agent, severe acute respiratory syndrome coronavirus 2 (SARS-CoV-2), is a novel betacoronavirus with phylogenetic similarity to SARS-CoV.^1^ To this date, in mid May 2020, the virus has infected over four million individuals worldwide and has claimed close to 300,000 lives. The mortality has been predominantly attributed to progressive pneumonia, leading to acute respiratory distress syndrome. Other organ failures and coagulopathy associated with COVID-19 has been observed in many studies, particularly in the hospitalized patients.^2–4^ The pandemic of COVID-19 overlaps with 2019–2020 flu season, which makes it difficult to differentially diagnose and manage patients with acute respiratory symptoms.

Current published clinical and laboratory data on COVID-19 are largely limited to studies with small sample size and mostly originated from China. Studies on temporal tracking of laboratory results in correlation with the severity of the disease are scarce. The most commonly reported laboratory abnormalities in COVID-19 include lymphopenia, prolonged prothrombin time (PT), and elevated lactate dehydrogenase (LDH).^3^ A study showed that a predictive model using laboratory values had a stronger discriminatory power predicting mortality in hospitalized patients compared to the clinical models.^5^ Laboratory manifestations in the western population and their correlation with the disease course of the hospitalized COVID-19 patients, especially their differences from those in influenza patients, are unclear.

In this study, we compared the laboratory manifestations of COVID-19 patients admitted to Northwestern Medicine Health System and compared these findings to a cohort of influenza patients. We explored laboratory parameters that could predict the mortality in the hospitalized patients.

## Methods

### Study design

We retrospectively collected demographic and clinical data from 973 patients with COVID-19 and 719 patients with influenza (including influenza A and B) from Northwestern Medicine Health System during the flu season of 2020 from January to April 2020. Most of the patients were adult over 20 years of age since the pediatric patients were managed at other institutions. Data collected include laboratory results from the day of presentation to day 14 of admission and clinical information until early May 2020. Many patients, especially those with influenza, did not have a full range of data sets since they were not hospitalized. We compared these laboratory results between patients with COVID-19 and influenza and identified the most significantly different parameters between these two diseases. We then selected 154 and 23 hospitalized COVID-19 and influenza patients, respectively, with at least 7 days of data points in each of these parameters and applied for a hierarchical clustering analysis. Five patients in the COVID-19 cohort were not grouped in any clusters, therefore excluded from further analyses. Patients in different clusters from the analysis were compared for their demographic information, including age and sex, underlying medical conditions, and clinical outcomes including fatality rate and length of hospital stay. The study protocol was approved by the institutional review board at Northwestern University. The requirement for informed consent was waived approved by the review board.

### SARS-CoV-2 detection

Samples were collected using a nasopharyngeal swab or by bronchoalveolar lavage (BAL). The swabs were then placed into a collection tube with 3 ml of virus preservation solution. For Real-time Reverse Transcriptase Polymerase Chain Reaction (RT-PCR) assay, total RNA was isolated using the QIAamp MinElute Virus Spin kit (Qiagen, Valencia, CA, USA). The RT-PCR assay was developed in the molecular diagnostic laboratory at Northwestern Memorial Hospital, using primers, probes, reagents and procedure designated by the Center for Disease Control and Prevention (CDC). The viral RNA was amplified using a one-step procedure using TaqPath 1-step RT qPCR master mix and CDC designed primers and probes (2019-nCOV Kit, IDT Coralville, IA). The products were amplified on the Quant Studio 6 flex system (Thermo Scientific, Waltham, MA, USA) or Xpert Xpress SARS-CoV-2 system (Cepheid, Sunnyvale, CA, USA).

### Influenza testing

Nasopharyngeal swabs were tested for influenza viruses with two assays: Xpert Xpress Flu/RSV from Cepheid Incorporation (Sunnyvale, CA, USA) for testing patients in the emergency room and FilArray Respiratory Panel 2 from BioMerieux (Marcy-l’Étoile, France) for inpatient testing. The Xpert Xpress Flu/RSV assay is a real-time RT-PCR-based assay for the detection and differentiation of influenza A and B viral RNA. The test was performed with the Cepheid GeneXpert system. The assay targets unique gene sequences that encode influenza A matrix (M), influenza A basic polymerase (PB2), and influenza A acidic (PA) proteins for influenza A virus and influenza B matrix (M) and influenza B nonstructural (NS) proteins for influenza B virus. The FilmArray Respiratory Panel 2 (RP2) test consists of automated nucleic acid extraction, reverse transcription, nucleic acid amplification, and result analysis in approximately 45 min per specimen.

### Data collection and statistical analysis

The clinical and laboratory information were extracted from electronic medical records of Northwestern Memorial Hospital. A two-tailed, unpaired t test was used to compare laboratory parameters between patients with COVID-19 and influenza. The heat map and clustering of 154 COVID-19 patients (at least 7 data points in the first 14 days of hospitalization) were analyzed by online Morpheus (Broad Institute, Boston, MA). Hierarchical clustering was performed with one minus spearman rank correlation with average linkage within patients. The clinical outcomes were compared among the identified clusters using logistic regression for binary outcomes (ICU use, intubation, discharged alive, and other respiratory infections) and Cox proportional hazards model for time-to-event outcomes (in-hospital death and death censored hospitalization stay). Because some of the events were rare, Firth’s bias reduction method based on penalized likelihood was used.^6^ The survival curve and death-censored hospital stay curve were estimated using Kaplan-Meier method, and the significance was assessed using the Log-rank test. p<0·05 was considered statistically significant. Statistical analyses were performed using the Prism software (version 8·0) and R (version 3·5·1).

## Results

We collected clinical and laboratory data from 973 patients with COVID-19 and 719 patients with influenza (including influenza A and B) from January to April 2020. Most of the patients were over 20 years of age. The influenza cohort included 274 males and 445 females with the diagnostic samples collected between January 1, 2020 and April 3, 2020. The COVID-19 cohort includes 507 males and 465 females with the samples collected between March 14, 2020 and April 21, 2020. We first compared the differences in the prevalence of the diseases in different age groups. Consistent with prior reports, influenza was more prevalent in younger patients^7^ whereas COVID-19 was more common in older patients^4,8^ (**Figure S1**). The fatality rate was higher in COVID-19 patients (35/973, 3·5%) than influenza patients (8/719, 1·1%). We evaluated clinical and laboratory results at presentation. Compared to influenza patients, patients with COVID-19 presented with significantly higher levels of red blood cell (RBC) count, hemoglobin, and platelet count. The level of lactate dehydrogenase (LDH) was significantly higher in patients with influenza. Troponin was higher in COVID-19 patients. The other laboratory parameters remained no statistically significant differences between influenza and COVID-19 (**Table S1**).

We compiled and temporally tracked all available laboratory results in these patients from the day of presentation to day 14 of the disease process. Overall, there were multiple parameters that showed significant differences between COVID-19 and influenza (**Figure 1**). In hematologic parameters, the white blood cell (WBC) count gradually increased in both diseases until day 8–9 when influenza patients started to decrease. However, WBC counts continued to increase in patients with COVID-19, which was mainly due to the increased absolute neutrophil count (ANC). The absolute lymphocyte count (ALC) in COVID-19 patients was stable at the borderline reference range during this period. In comparison, influenza patients showed more fluctuating lymphocyte counts. Hemoglobin and RBC in both groups gradually reduced over time. However, COVID-19 patients dropped more rapidly and plummeted to the same levels as influenza patients in the later stages of disease course. The platelet counts were significantly higher in COVID-19 patients, although thrombocytopenia was rare during this clinical course in both diseases (**Figure 1A**).

**Figure 1:**
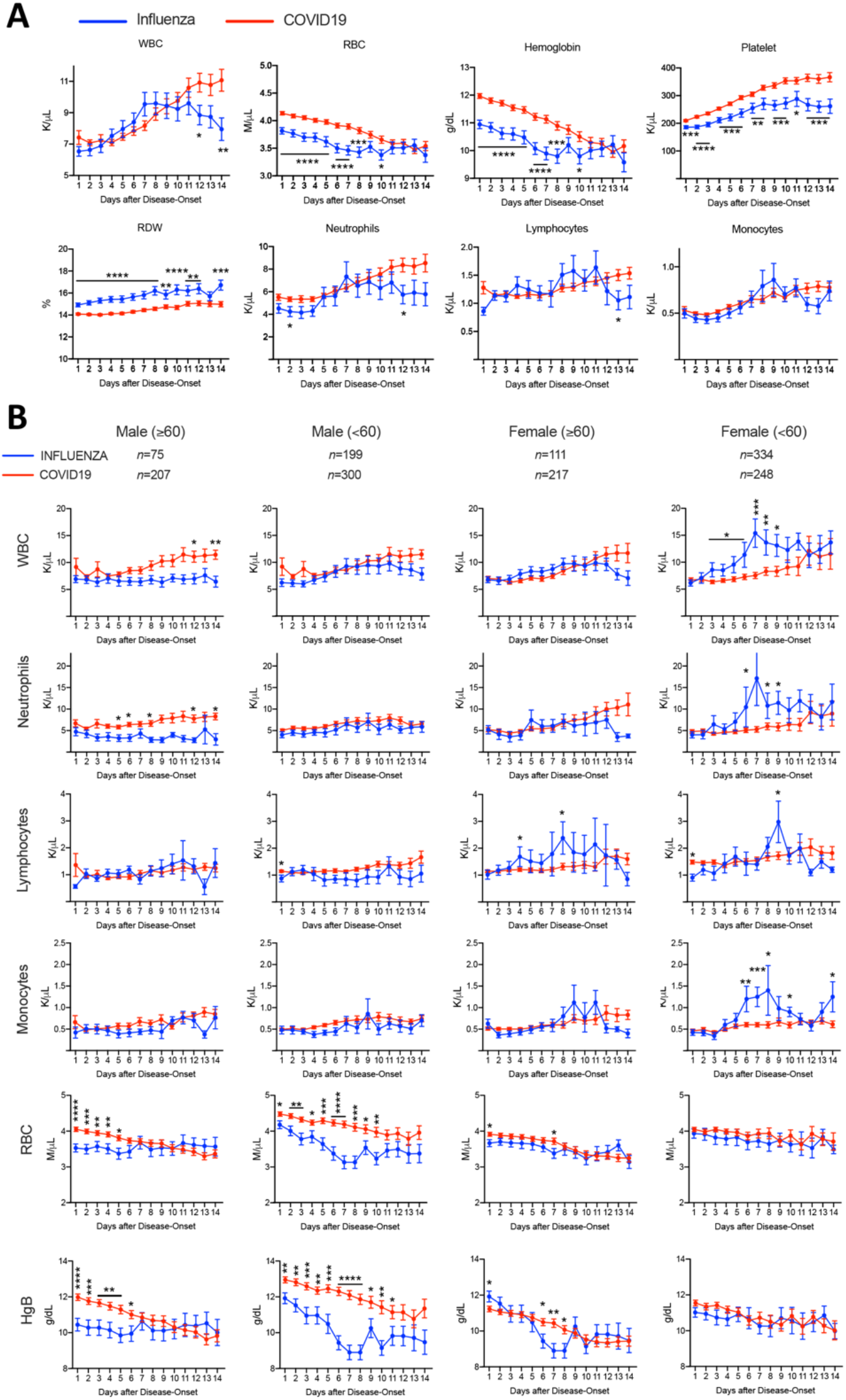
**Comparison of Hematology Laboratory Results between Influenza and COVID-19 patients.** Complete blood counts were collected from admission to day 14 of hospitalization, and presented as Mean ± SEM within timeline charts in A. The patients were further grouped based on age and gender and analyzed in B. WBC: whole blood counts. RBC: red blood cells. RDW: red blood cell distribution width. HgB: Hemoglobin. *p<0·05, **p<0·01, *** p<0·001, ****p<0·0001

We separated the patients in each group based on sex and age (<60 or ≥60) (**Figure 1B**). WBC increased in all subgroups of COVID-19 patients with the increase most significant in older (≥60 years old) male patients. The same was observed for ANC. Interestingly, WBC in younger (< 60 years old) female influenza patients increased significantly in the early stages of disease course followed by a gradual decrease, which could be seen in most of the leukocyte lineages. More important, the differences in the level of anemia were primarily contributed by male COVID-19 patients. In addition, older male patients were more anemic at presentation and during the course.

Serum biochemical tests also revealed statistically significant differences between COVID-19 and influenza (**Figure 2**). Alanine aminotransferase (ALT) and blood urea nitrogen (BUN) were started at around the higher reference range in patients with both diseases. While their levels remained relatively constant in patients with influenza, ALT and BUN gradually increased and became significantly higher in the late stages of disease course in COVID-19 patients. This was most prominent in male patients whereas the differences were not significant in female patients (**Figure 2A**). The level of D-dimer started at increased levels in both diseases. The level continued to climb in patients with COVID-19 in both males and females, although there was no statistical significance due to inadequate data points in influenza patients (**Figure 2B and S2A**). The levels of C-reactive protein (CRP), ferritin, and fibrinogen in COVID-19 patients were significantly increased and reached the highest level at 6–8 days, which were followed by gradual decrease (**Figure 2B**). Notably, clinical data of these parameters in influenza patients were not as comprehensive as those in the COVID-19 patients, which compromised statistical analyses when divided based on age and sex. LDH was significantly higher in influenza patients (**Figure 2C**). The levels of alkaline phosphatase and bilirubin were slightly higher in influenza patients than COVID-19 patients over the 14-day course, although there was no statistical significance. The other parameters tested, including aspartate transaminase (AST), creatinine, prothrombin time (PT), partial thromboplastin time (PTT), and troponin, did not show differences between influenza and COVID-19 groups (**Figure S2B**).

**Figure 2:**
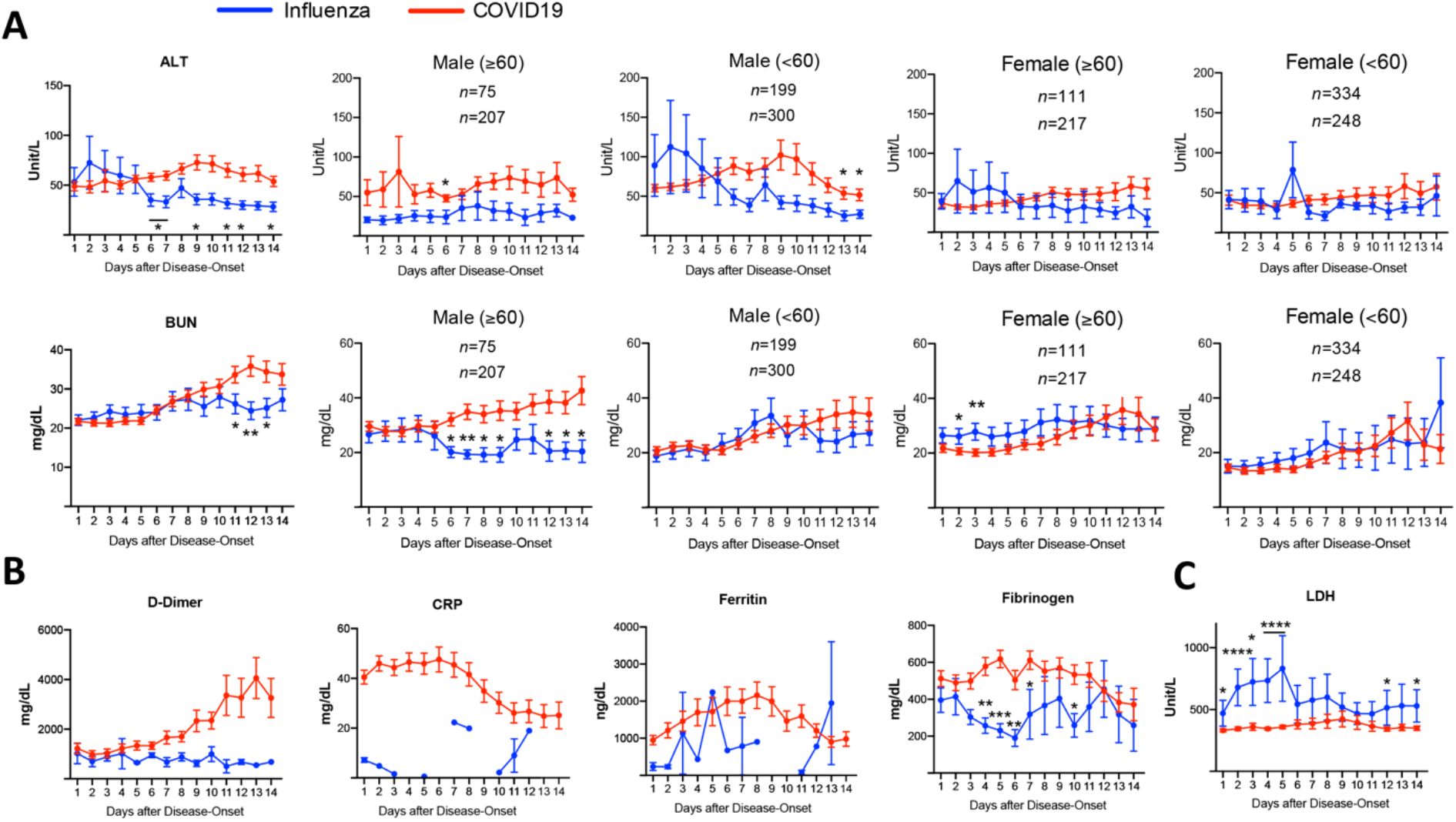
**Changes of Serum Biochemical Results in Influenza and COVID19 Groups Over the 14-day Hospitalization.** ALT: alanine aminotransferase. BUN: blood urea nitrogen. CRP: C-reactive protein. LDH: lactate dehydrogenase. *p<0·05, **p<0·01.

These analyses reveal that overall, COVID-19 patients tend to have higher hemoglobin and red blood cell counts but more rapid decline, higher white blood cell and neutrophil counts especially in the late stages of disease course, and higher ALT, BUN, and D-Dimer. However, it is unclear whether there are any correlations among these parameters in COVID-19 patients, and whether there are specific groups in these patients that may show distinct clinical outcomes. To answer these questions, we performed a hierarchical clustering analysis to compare various laboratory parameters in individual patient over the 14-day course. For the laboratory parameters, we focused on complete blood count, D-dimer, BUN, and ALT since they were significantly different between patients with influenza and COVID-19. We chose 154 COVID-19 and 23 influenza inpatients with at least 7 days of data points in each of these parameters to analyze. Five clusters (1 to 5 from far left to far right, respectively) were revealed in COVID-19 patients whereas no distinct clusters were identified in influenza patients due to lack of sufficient patients (**Figure 3**). Cluster 1 showed severe worsening anemia, increased red blood cell distribution width (RDW), leukocytosis (including neutrophilia and monocytosis), higher BUN, creatinine, D-dimer, alkaline phosphatase, bilirubin, and troponin (**Figure 4A**). These parameters were much milder in other clusters, especially in clusters 3 and 4. The level of CRP was high in cluster 2 (**Figure 4B**). Other parameters, including lymphocyte and platelet counts (**Figure 4A**), ALT, fibrinogen, ferritin, PT, PTT, LDH, oxygen saturation, and maximum temperature remained no differences among these clusters (**Figure S3**).

**Figure 3:**
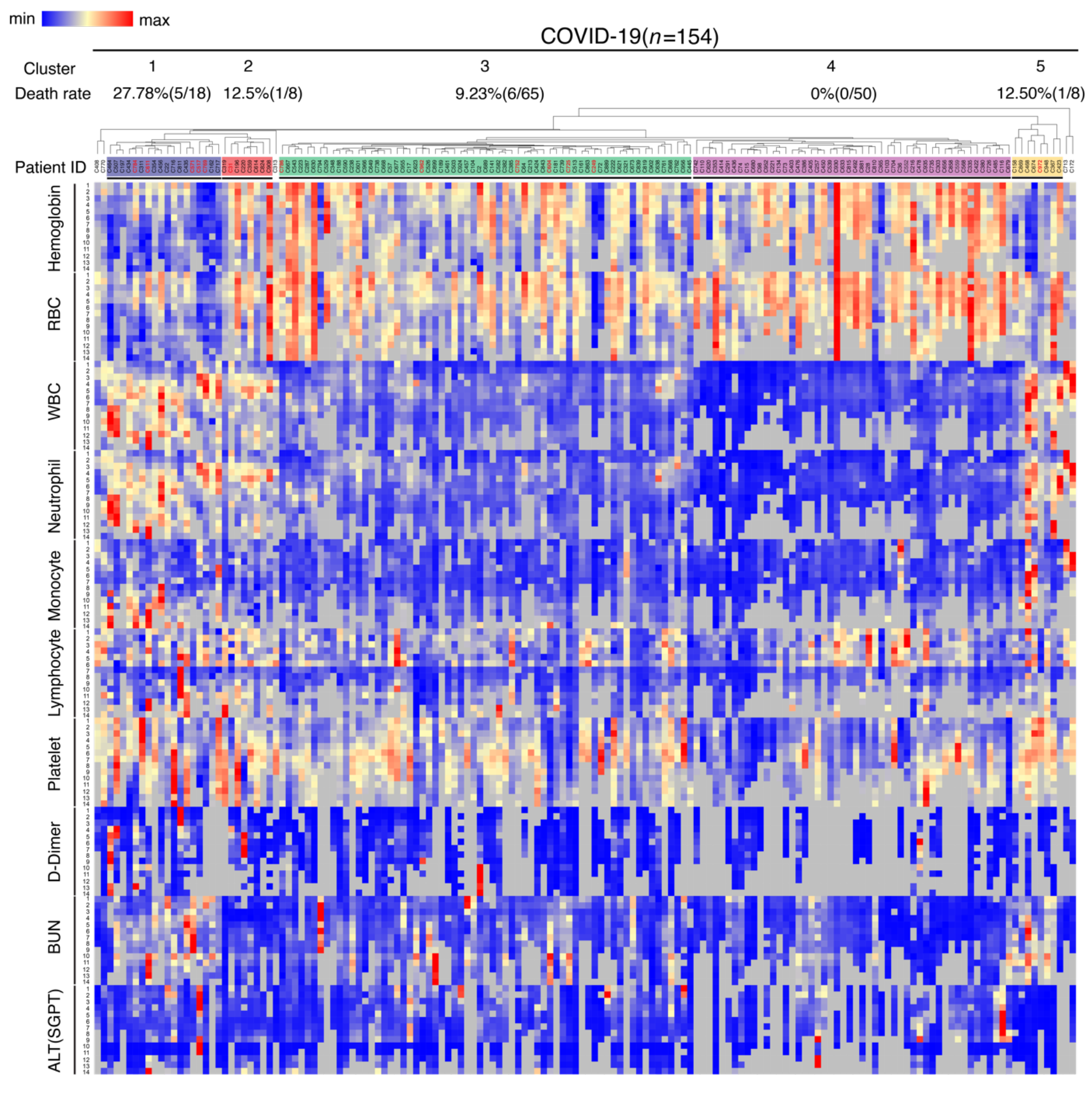
**Hierarchical Clustering analysis of COVID-19 cohort.** Patient IDs in red represent deceased patients during hospitalization.

**Figure 4:**
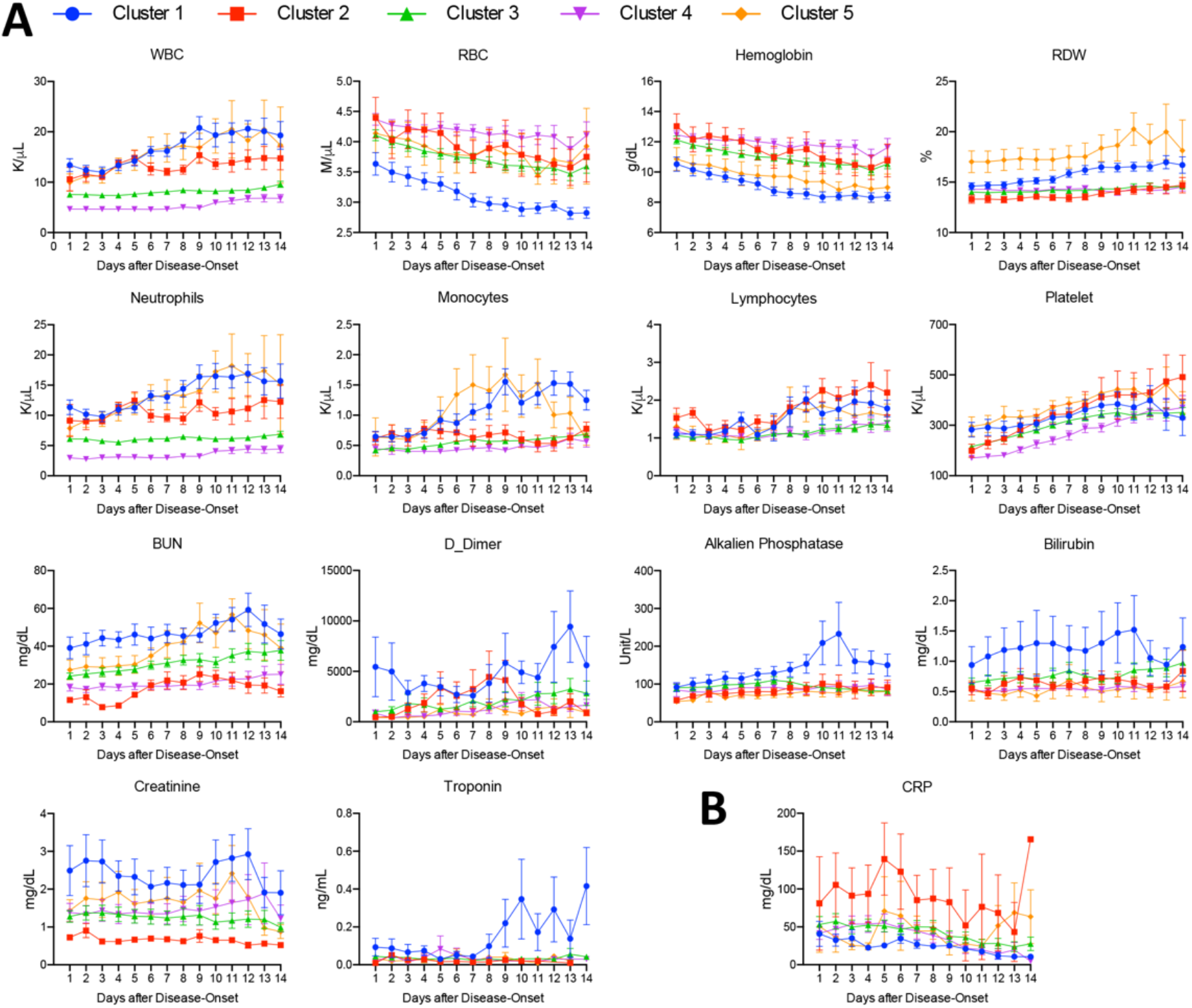
**Comparison of Laboratory Results over the 14-day Hospitalization among COVID-19 Clusters.**

We further investigated the clinical manifestation of patients in these different COVID-19 clusters. Cluster 1 showed the highest mortality rate (27·8%), which was followed by group 2 and 5 (12·5%). Group 3 and 4 had the lowest mortality rate (9% and 0%, respectively) (**Table S2 and Figure 5A**). There were also significantly increased rates of ICU admission (odds ratio 18·85, 95% CI 4·20–180·23), intubation (odds ratio 40·07, 95% CI 8·62–391·26), and other respiratory infections (odds ratio 26·10, 95% CI 6·79–127·49) in cluster 1 compared to cluster 4 (**Table S3**). Notably, there was no significant differences in age distributions in these clusters although males were predominant in most of the clusters, including clusters 3 and 4. The majority of the patients in all clusters had pre-existing conditions, mostly hypertension and type 2 diabetes (**Table S4**). Compared to cluster 4, the other groups showed significantly increased length of hospital stay, especially for cluster 1 (**Figure 5B**).

**Figure 5:**
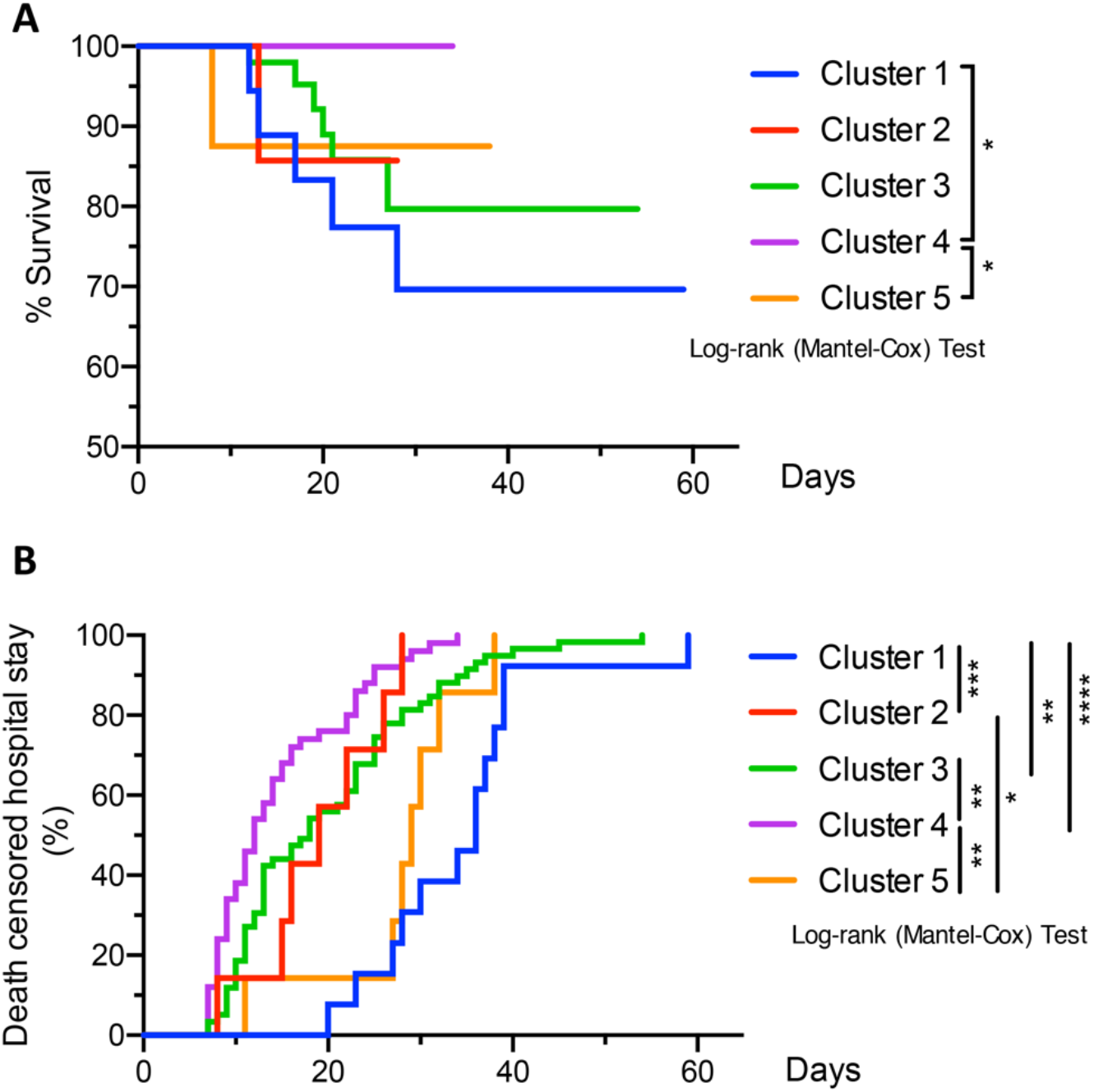
**Analyses of Survival and Death-censored Hospitalization Rates of Different Clusters in COVID-19 Cohort.** *p<0·05, **p<0·01, *** p<0·001, ****p<0·0001

## Discussion

The overlap of flu season with the pandemic of COVID-19 complicates the clinical management of patients with respiratory symptoms. Studies of direct comparison of clinical and laboratory results between these two infectious diseases are rare. A recent report compared hospitalized patients with acute respiratory distress syndrome (ARDS) caused by COVID-19 and influenza H1N1. The authors concluded that ARDS patients with COVID-19 had lower severity of illness at presentation and lower mortality adjusted by sequential organ failure assessment scores^9^. However, the study used data from two independent institute, which could lead to certain bias. It is also unclear the differences on the clinical and laboratory results during the hospitalization of these patients. To address this question, our study showed that there were significant differences between hospitalized patients with COVID-19 and influenza when we followed their temporal changes of laboratory results. Compared to influenza patients, the most significant differences over the course of 14 days of hospitalization in COVID-19 patients were worsening anemia, worsening leukocytosis, and an increase in D-dimer, BUN, and ALT.

Instead of comparing clinical endpoints to evaluate risks as performed in most of the published studies, we stratified the hospitalized COVID-19 patients through clustering of their laboratory results that were most significantly different from influenza patients (i.e. complete blood count, D-dimer, BUN, and ALT) during the first 14 days of hospitalization. The major differences in these clusters were red cell indices, neutrophil count, monocyte count, and BUN. Surprisingly, lymphocyte count did not show differences as prominent as the other parameters among these patients. Lymphopenia was shown to be common and correlated with a worse prognosis in COVID-19 patients.^10^ A recent meta-analysis showed a 3-fold higher risk of developing severe COVID-19.^11^ However, the studies involved in this meta-analysis were all data from patients in China. The findings in our cohort may represent distinctive features in the western population. In fact, the majority of the COVID-19 patients in our study showed lymphocyte counts within the reference range over the 14-day hospitalization (low level reference 1·0 × 10^9^/L), even in patients in cluster 1 with the worst prognosis.

Reviewing of the clinical record, including the record after the initial 14 days, revealed that cluster 1 had the worst outcomes including high fatality, high ICU admission and intubation rate, and prolonged hospital stay. Compared to cluster 4 with the best outcome, patients in cluster 1 showed features consistent with previously published reports, including high WBC, BUN, creatinine, alkaline phosphatase, bilirubin, and troponin.^4,5,8,12^ However, severe anemia was not reported as a prognostic feature. A recent report showed no differences in hemoglobin between survival and non-survival COVID-19 patients in China.^13^ Therefore, worsening anemia represents another distinctive feature of worse prognosis in our cohort, especially for male patients. Notably, anemia was associated with neutrophilia and monocytosis in cluster 1, which could also be related to additional respiratory infections that were more frequent in these patients. These laboratory features associated with a worse prognosis were more prominent in males. Indeed, of the 13 deceased patient in all clusters, 10 were males. The majority of the patients in all 5 clusters had more than 1 comorbidity, which is not a confounding factor in risk stratification in this study.

Our hierarchal clustering approach is unique in a way that the pathophysiology of COVID-19 could be indicated by laboratory values over time. For example, cluster 1 may represent patients who were showing a clotting/vascular system pathology with multi-organ dysfunction (elevated D-dimer, and abnormal cardiac, renal, and liver function tests). Cluster 2 may represent cytokine storm or secondary bacterial pneumonia (elevated CRP) without involvement of other organs. Further studies on specific organ systems may be useful to confirm the underlying pathology in patients in these clusters. The clustering approach may also be useful to identify patients responsive to specific therapies.

Some other clinical and laboratory results that were reported to be distinctive between mild and severe COVID-19 patients were not informative when we evaluate the risks in our cohort. There were no differences in daily minimum oxygen saturation or maximum temperature among the clusters. In the laboratory data, there were no difference in ALT, fibrinogen, PT, PTT, ferritin and LDH over the 14-day course. These laboratory parameters could be important in differentiating hospitalized and non-hospitalized patients, which is not the focus of this study.

Our study has its limitations. First, the patients were limited to a single healthcare system in Chicago metropolitan area. Second, due to the lack of sufficient number of influenza patients with over 7 days of laboratory data sets in our hierarchical analysis, we could not perform the same risk stratification in these patients. It remains to be determined whether similar laboratory patterns as in COVID-19 cluster 1 are also present in severe hospitalized influenza patients. Third, because of the same reason, several laboratory parameters did not show statistical significance between COVID-19 and influenza, although they appeared to be apparently different. Overall, our findings provide values to predict risk groups for further management in hospitalized COVID-19 patients in the western population. Further prospective studies using independent groups will be informative to confirm these findings.

## Data Availability

N/A

## Reference

1. Zhu N, Zhang D, Wang W, et al. A Novel Coronavirus from Patients with Pneumonia in China, 2019. N Engl J Med 2020; 382(8): 727–33.

2. Tang N, Li D, Wang X, Sun Z. Abnormal coagulation parameters are associated with poor prognosis in patients with novel coronavirus pneumonia. J Thromb Haemost 2020; 18(4): 844–7.

3. Wang D, Hu B, Hu C, et al. Clinical Characteristics of 138 Hospitalized Patients With 2019 Novel Coronavirus-Infected Pneumonia in Wuhan, China. JAMA 2020.

4. Zhou F, Yu T, Du R, et al. Clinical course and risk factors for mortality of adult inpatients with COVID-19 in Wuhan, China: a retrospective cohort study. Lancet 2020; 395(10229): 1054–62.

5. Wang K, Zuo P, Liu Y, et al. Clinical and laboratory predictors of in-hospital mortality in patients with COVID-19: a cohort study in Wuhan, China. Clin Infect Dis 2020.

6. Firth D. Bias reduction of maximum likelihood estimates. Biometrika 1993; 80(1): 12.

7. Tokars JI, Olsen SJ, Reed C. Seasonal Incidence of Symptomatic Influenza in the United States. Clin Infect Dis 2018; 66(10): 1511–8.

8. Bhatraju PK, Ghassemieh BJ, Nichols M, et al. Covid-19 in Critically Ill Patients in the Seattle Region – Case Series. N Engl J Med 2020.

9. Tang X, Du R, Wang R, et al. Comparison of Hospitalized Patients With ARDS Caused by COVID-19 and H1N1. Chest 2020.

10. Tan L, Wang Q, Zhang D, et al. Lymphopenia predicts disease severity of COVID-19: a descriptive and predictive study. Signal Transduct Target Ther 2020; 5(1): 33.

11. Zhao Q, Meng M, Kumar R, et al. Lymphopenia is associated with severe coronavirus disease 2019 (COVID-19) infections: A systemic review and meta-analysis. Int J Infect Dis 2020.

12. Rodriguez-Morales AJ, Cardona-Ospina JA, Gutierrez-Ocampo E, et al. Clinical, laboratory and imaging features of COVID-19: A systematic review and meta-analysis. Travel Med Infect Dis 2020: 101623.

13. Ruan Q, Yang K, Wang W, Jiang L, Song J. Clinical predictors of mortality due to COVID-19 based on an analysis of data of 150 patients from Wuhan, China. Intensive Care Med 2020.

